# Functional consequences of genetic risk for neuropsychiatric conditions at chr22q

**DOI:** 10.1101/2025.07.01.25330642

**Authors:** Ajay Nadig, Matthew Tegtmeyer, Ajinkya Patil, Yingzhe Zhang, Emi Ling, Daniel Weiner, Dhara Liyanage, Serkan Erdin, Rachita Yadav, Jonas Bybjerg-Grauholm, Ryan L Collins, iPSYCH Consortium, Psychiatric Genomics Consortium Autism Working Group, Jakob Grove, Anders D. Børglum, Michael Talkowski, Luke O’Connor, Cigall Kadoch, Steven McCarroll, Ralda Nehme, Elise B. Robinson

## Abstract

Understanding how common and rare genetic variation raises risk for neuropsychiatric disease remains a major challenge. We identify the long arm of chromosome 22 (chr22q) as a region where common polygenic risk for schizophrenia, autism, ADHD, and lower IQ is associated with coordinated downregulation of gene expression in postmortem human brain tissue. The effects are strikingly consistent between neuropsychiatric diagnoses and across brain cell types, and appear to be specific to brain-related traits. We observe that common variant risk for neuropsychiatric diseases has remarkably diffuse expression associations across chr22q, including long-range aggregate associations between genetic variants and genes over 10 Mb away. Polygenic risk for psychiatric disease at chr22q is more strongly associated with lower cognitive ability than elsewhere in the genome, suggesting phenotypic convergence with the 22q11.2 deletion, a rare genetic disorder that causes intellectual disability, schizophrenia, autism, and ADHD. Using human iPSC data, we show that the 22q11.2del induces similarly broad expression downregulation across chr22q in multiple neural cell types and experimental settings. Altogether, our results nominate chr22q as a regulatory hub in neuropsychiatric disease, where common and rare genetic risk factors converge both functionally and phenotypically.

## Introduction

Over the last decade, genetic studies have yielded hundreds of reproducible associations to neuropsychiatric disorders. These associations include rare, single nucleotide and copy-number variants of large effect (Singh et al. 2022; Marshall et al. 2017; Fu et al. 2022), as well as many thousands of common variants of small effect that – in aggregate – explain the majority of neuropsychiatric disease heritability (Loh et al. 2015; Trubetskoy et al. 2022). The “variant-to-function” (V2F) problem is indeed our field’s next hurdle. We must select disease-associated variants on which to focus, and connect them to coherent and targetable biological processes. With long association lists in hand, genes and processes that have been nominated by multiple genetic associations – e.g. by both rare and common variants – are particularly strong candidates for study.

We recently described a new type of genomic convergence, one in which two of autism’s strongest genetic predictors – 1) common polygenic variation and 2) deletions at 16p11.2 – cause widespread and correlated decreases in gene expression across the ∼30 Mb p arm of Chromosome 16 (16p; Weiner et al. 2022). In addition to highlighting an autism-relevant genomic region through multiple classes of genetic variation, this finding nominated a new mechanistic hypothesis: that subtle functional changes across large genomic territories could be particularly relevant to risk for neuropsychiatric disorders. Patterning of the gene expression consequences on chr16p led us to speculate involvement of the chromosome arm’s unusually dense intrachromosomal physical contact, which is a function of the genome’s folding patterns. Folding allows the genome to fit into individual cells, and its patterning renders some genes, like those within chr16p, in greater 3D contact than expected given the linear distance between them (Rao et al. 2014).

In this analysis, we focus on the q arm of Chromosome 22 (chr22q), having observed that it is one of very few genomic territories with density of physical contact similar to that of chr16p (**Figure 1**). This similarity is remarkable, as chr22q also harbors multiple CNVs mediated by a mechanism that involves mispairing of a cluster of large, duplicated sequences spanning chr22q11.2 (Shaikh et al. 2000). These chr22q and chr16p CNVs collectively represent two of the most common recurrent genomic disorder loci in the human genome that are associated with a spectrum of developmental disorders and neuropsychiatric disease (Malhotra and Sebat 2012). Indeed, 22q11.2del is enormously variable in its impact, but confers strong risk for schizophrenia, intellectual disability, and autism, along with a constellation of other medical concerns including cardiac malformation and immunodeficiency (McDonald-McGinn et al. 2015). While the cardiac manifestations of 22q11.2del have been associated with a specific gene in the deletion interval, *TBX1* (Yagi et al. 2003), the deletion’s association to neuropsychiatric disease remains poorly understood (but see also Nehme et al. 2022; Khan et al. 2020).

**Figure 1:**
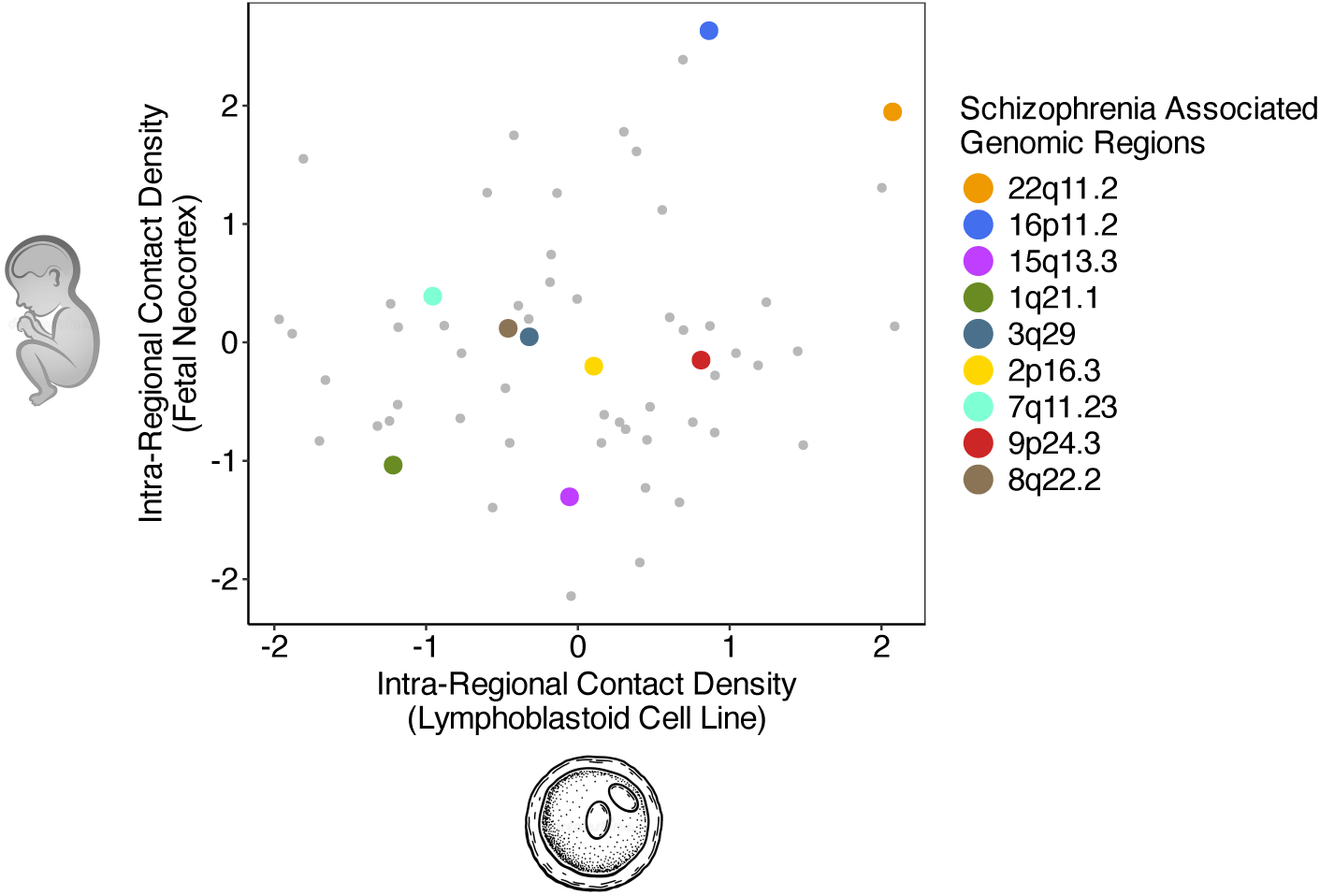
Intra-chromosomal contact in large genomic territories. Degree of chromosomal contact within 33-Mb regions tiling the human genome (**Methods**) from lymphoblastoid cell lines (Lieberman-Aiden et al. 2009) and fetal neocortex (Won et al. 2016). Regions containing CNVs associated with schizophrenia and other neuropsychiatric diseases (Marshall et al. 2017) are shown in color.

The majority of genetic risk for neuropsychiatric disorders is *common and polygenic*, comprising many thousands of variants spread evenly across the genome, operating in aggregate and carried by everyone to some degree. In this work, we integrate several new resources to characterize the molecular consequences of diverse, disease-associated genetic variation in chr22q, both common and rare. We provide further evidence that uncommonly diffuse transcriptomic impact, inflated within contact-dense regions of the genome, may have a special relationship to the biology of neuropsychiatric disease. Using postmortem human brain tissue, we find that common polygenic risk for several neuropsychiatric diagnoses *decreases* the average expression of hundreds of genes on chr22q. The gene expression consequences are shared to an extraordinary degree among the common variant risk factors for schizophrenia, intellectual disability, autism, and ADHD. Polygenic risk for diagnoses unrelated to the brain (such as height, inflammatory bowel disease), does not manifest this property. Using new analysis approaches, we find evidence that the polygenic expression consequences arise from previously uncharacterized, supra-megabase V2F associations, which, like in chr16p, may relate to chr22q’s exceptional 3D architecture. Next, we demonstrate that the 22q11.2del, on average, results in similar chromosome-arm-wide decreases of gene expression *in vitro*. Lastly, by aggregating data from multiple 22q11.2del *in vitro* experiments, we quantify experimental variability in the transcriptomic consequences of 22q11.2del, highlighting the extent to which large datasets, meta-analysis, and broad collaboration will be needed to reliably and reproducibly interrogate the functional consequences of disease-associated genetic variation.

## Results

### Within chr22q, polygenic risk for neuropsychiatric disease consistently downregulates gene expression

Common polygenic variation poses challenges for traditional approaches to functional interrogation: there a large number of associated variants, each of modest effect size (Trubetskoy et al. 2022; Loh et al. 2015; O’Connor 2021), and often cryptic associations between those variants and genes (Gazal et al. 2022; Weissbrod et al. 2020; Wu et al. 2023).

To interrogate the transcriptomic consequences of common polygenic risk for neuropsychiatric disease within chr22q, we developed approaches to aggregate signals across variants rather than examining each locus individually. We first estimated associations between: i) expression of 392 genes within chr22q, estimated at single cell resolution in postmortem brain tissue from 122 donors (Ling et al. 2024), and ii) polygenic risk for multiple disease outcomes, specifically within chr22q (from here: local polygenic risk; **Methods**). We used LDPred2-inf (Privé, Arbel, and Vilhjálmsson 2021) to compute local polygenic scores at chr22q for 4 neuropsychiatric phenotypes – schizophrenia, autism, ADHD, and lower IQ – each of which is associated with the 22q11.2del (Olsen et al. 2018; Schneider et al. 2014). We also estimated local polygenic risk for 6 non-neuropsychiatric control traits (body mass index, height, serum LDL, red blood cell count, allergic conditions, and irritable bowel disease; **Methods**). We found that local polygenic risk for each of the neuropsychiatric conditions was associated with a small decrease in average expression across chr22q genes, in glutamatergic neurons (**Figure 2A; Methods**), as well as GABAergic neurons and astrocytes, but not for oligodendrocytes (**Supplementary Figure 1**).

**Figure 2:**
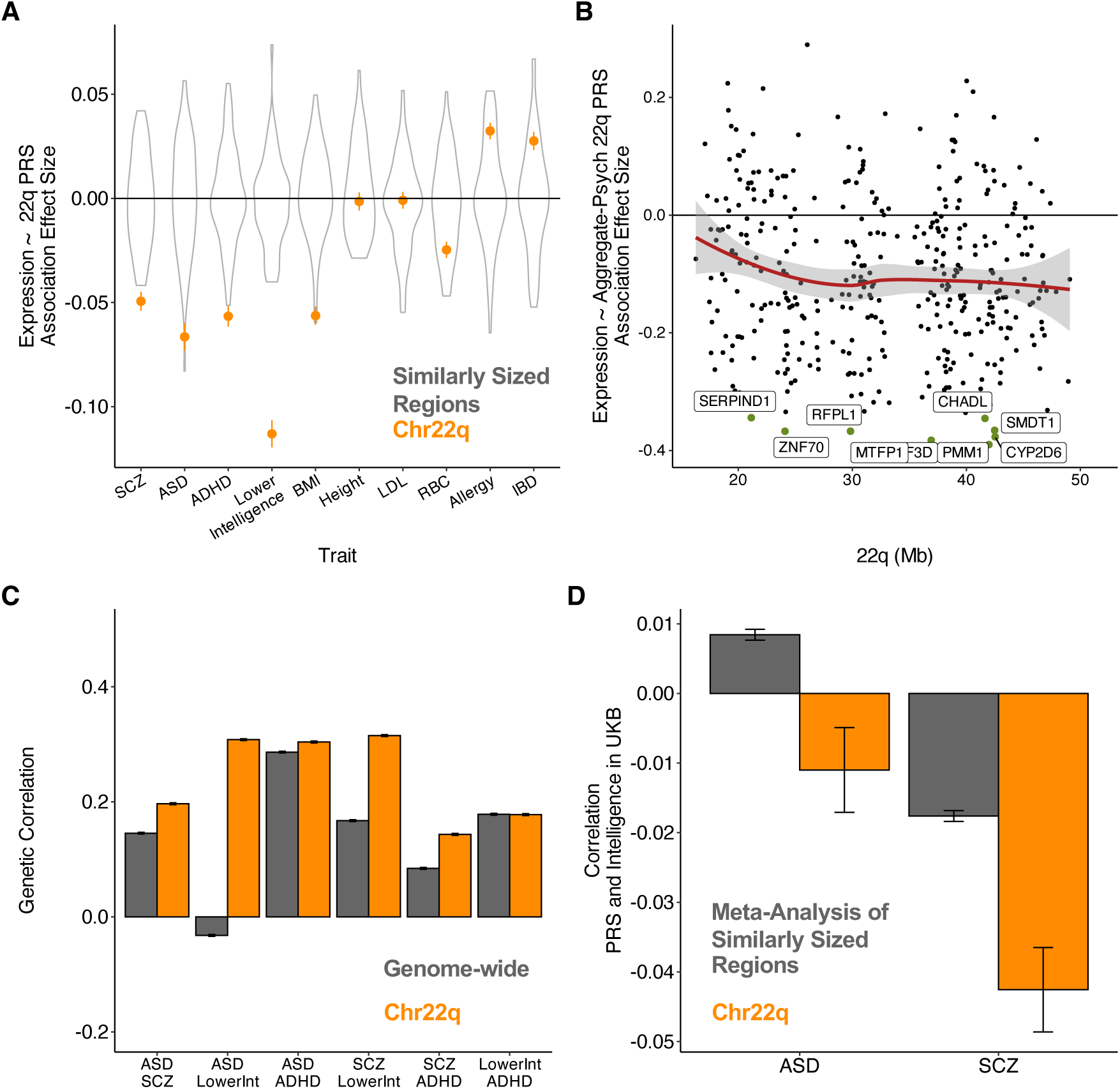
Association between local polygenic risk for neuropsychiatric disorders and lower gene expression at chr22q. (A) Associations between local polygenic scores for various diagnoses and traits and average gene expression in glutamatergic neurons at chr22q (orange) and 60 other similarly sized genomic territories (gray). (B) Gene-wise associations between chr22q local polygenic score, aggregated across neuropsychiatric diagnoses/traits, and chr22q gene expression. Labels indicate genes that have Bonferroni-significant associations. (C) Inter-diagnosis genome-wide (gray) and local chr22q (orange) genetic correlations, estimated by correlating polygenic risk scores in the UK Biobank. (D) Associations between the Aggregate-Psych polygenic score and fluid intelligence score in the UK Biobank, shown for chr22q (orange) and a meta-analysis of 60 similarly sized genomic regions (gray).

To assess the extent to which this observation was unique to chr22q, we computed similar local polygenic risk scores for 60 other genomic regions of the same size (**Methods**). We found that the association between polygenic risk for neuropsychiatric diagnoses/traits and decreased average gene expression was largest at chr22q (**Figure 2A**). In our analyses of non-neuropsychiatric traits, we observed a similar mean expression downregulation for body mass index (BMI; p < 0.001), but no mean expression association with polygenic scores for height (p = 0.37) or LDL (p = 0.4), a mean expression upregulation for allergy (p < 0.001) and inflammatory bowel disease (IBD; p < 0.001), and a smaller mean expression down regulation for red blood cell count (p < 0.001) (**Figure 2A**). This suggests some specificity for neuropsychiatric traits, at least for the brain cell types examined. The unusual finding for BMI likely owes to the strong enrichment of BMI heritability in genes expressed in the central nervous system (Finucane et al. 2018; Loos and Yeo 2022). While chr22q is indeed uncommonly gene dense, and further enriched for genes expressed in brain tissues, those characteristics cannot explain its exceptional transcriptional effects (**Methods**).

After observing an association between polygenic risk for all four neuropsychiatric traits and average gene expression within chr22q, we compared gene expression consequences between the traits. The common polygenic influences on SCZ, autism, ADHD and low IQ have significantly correlated, and directionally consistent, effects on gene expression – at the per gene level – within chr22q (0.31 to 0.68; **Supplementary Figure 2**). These correlations suggest remarkable similarity in the functional consequences of genetic risk for diverse neuropsychiatric diagnoses within chr22q, a degree of sharing made particularly surprising by the diagnoses’ genome-wide genetic relationships. Genome-wide genetic relationships are typically estimated through genetic correlations, which indicate the proportion of total common genetic influences shared between two traits (Lee et al. 2019). A genetic correlation of .2, for example, suggests that a pair of traits share 20% of their genome-wide genetic influences. The neuropsychiatric diagnoses included here share genetic correlations that range from modest and significantly negative (autism/lower intelligence, rg = –0.2; Grove et al, 2019) to moderate and significantly positive (autism/attention-deficit hyperactivity disorder, rg = 0.36; Grove et al, 2019). Their transcriptomic consequences within chr22q, on the other hand, are uniformly positive and larger in magnitude.

Given the degree of transcriptomic sharing, we averaged the four neuropsychiatric polygenic scores to boost power, inspired by a recent study showing that aggregating polygenic scores across traits can substantially improve trait prediction (Albiñana et al. 2023). We found that the resulting “Aggregate-Psych” polygenic score was associated with decreased expression of chr22q genes to a greater extent than our analysis of individual traits (p < 0.001; **Figure 2B**). This aggregated score also had sufficient power to resolve individual genes where expression was significantly associated with polygenic risk for neuropsychiatric disease, including *EIF3D* (p < 0.001), which is a gene known to be intolerant to loss-of-function (LoF) variation (observed / expected LoF variation in gnomAD = 0.11; pLI = 1) (Karczewski et al. 2020).

We explored the consistency of these observations in bulk post-mortem samples from the CommonMind consortium (**Supplementary Figure 3; Methods**). We found that the Aggregate-Psych polygenic score was associated with decreased bulk expression of chr22q genes in 328 European ancestry samples (p < 0.001), but not 206 African ancestry samples (p = 0.30). The failure of replication in samples of African ancestry likely reflects the fact that the polygenic score weights were derived from European-ancestry samples, and cross-population polygenic score performance is often poor (Martin et al. 2019).

### Phenotypic and functional convergence of neuropsychiatric disease risk within chr22q

In the analyses above, we observed local polygenic risk for each of schizophrenia, autism, ADHD, and lower intelligence to decrease gene expression at chr22q, and observed the similarity of these effects across traits to be larger than expected. For example, the correlation of transcriptomic responses to autism and lower intelligence PRS in glutamatergic neurons was large and positive on chr22q (0.68, **Supplementary Figure 2**). This seems inconsistent with the genome-wide genetic correlation between the two, which is negative (–0.20; (Martin et al. 2019; Grove et al. 2019). Put differently, while autism and lower intelligence have a negative genome-wide correlation – meaning that, in general, genetic risk factors for one tend to be protective for the other – their effects on gene expression at chr22q tell a different story.

One explanation for these findings may be that genetic risk factors for neuropsychiatric conditions are unusually similar on chr22q. Genetic correlations capture the *average* genetic relationship between two traits genome-wide, but trait relationships can indeed vary between regions of the genome (Shi et al. 2017; Werme et al. 2022). We estimated the genome-wide (including chr22q) and chr22q-specific PRS correlations across neuropsychiatric diagnoses/traits (**Figure 2C**). We found that local chr22q polygenic score correlations between schizophrenia/autism and lower intelligence were substantially higher, and more consistent in direction, than the corresponding genome-wide polygenic score correlations (autism/lower intelligence: local correlation = 0.31, se = 0.002, genome-wide correlation = –0.03, se = 0.002, comparison p < 0.001; schizophrenia/lower intelligence: local correlation = 0.32, se = 0.002, genome-wide correlation = 0.17, se = 0.002, comparison p < 0.001). Using a different estimation procedure, rho-HESS (Shi et al. 2017; Methods), we found a similar pattern of results, where neuropsychiatric diagnoses had higher genetic correlations with lower intelligence at chr22q (**Supplementary Figure 4**).

We next asked whether this unexpected pattern of polygenic risk sharing within chr22q has unexpected phenotypic consequences. If genetic risk at chr22q exhibits an unusual pattern of overlap across neuropsychiatric traits, genetic risk for neuropsychiatric disease may influence phenotypic outcomes in ways that differ from most regions of the genomic. To assess this question, we computed associations between chr22q polygenic scores and intelligence as estimated in the UK Biobank (**Methods; Figure 2D**). We compared association estimates from chr22q with meta-analyzed estimates of 60 similarly sized genomic regions (to avoid confounding by variable attenuation bias). We found that associations between local polygenic scores at chr22q and intelligence were significantly more negative than elsewhere in the genome for autism (comparison p = 0.001) schizophrenia (comparison p < 0.001). Notably, we observed a change in sign for autism, with the chr22q autism polygenic score being associated with lower intelligence. This result suggests a parallel between the phenotypic effects common, polygenic risk for neuropsychiatric disease at chr22q and those of the 22q11.2del, which is associated with risk for intellectual disability and neuropsychiatric diagnoses (Olsen et al. 2018); (Chawner et al. 2017).

### Common polygenic risk for neuropsychiatric diagnoses has uncommonly distal functional consequences within 22q

The above analyses demonstrate that, within chr22q, polygenic risk for multiple neuropsychiatric conditions is associated with an average decrease in gene expression. What induces this highly coordinated impact across a large genomic territory? We considered two possibilities (**Figure 3A**). Hypothesis 1: the relationship between local polygenic risk at chr22q and decreased gene expression is mediated by a series of short-range regulatory interactions between variants and their nearby genes. Hypothesis 2: there exist longer-range interactions where many variants exert convergent regulatory effects on faraway genes. Hypothesis 1 reflects contemporary understanding of expression quantitative trait loci (eQTL) that generally act in cis to regulate nearby genes; Hypothesis 2 reflects an alternative, unknown mechanism where common regulatory variants influence the expression of faraway genes across a chromosome arm.

**Figure 3.**
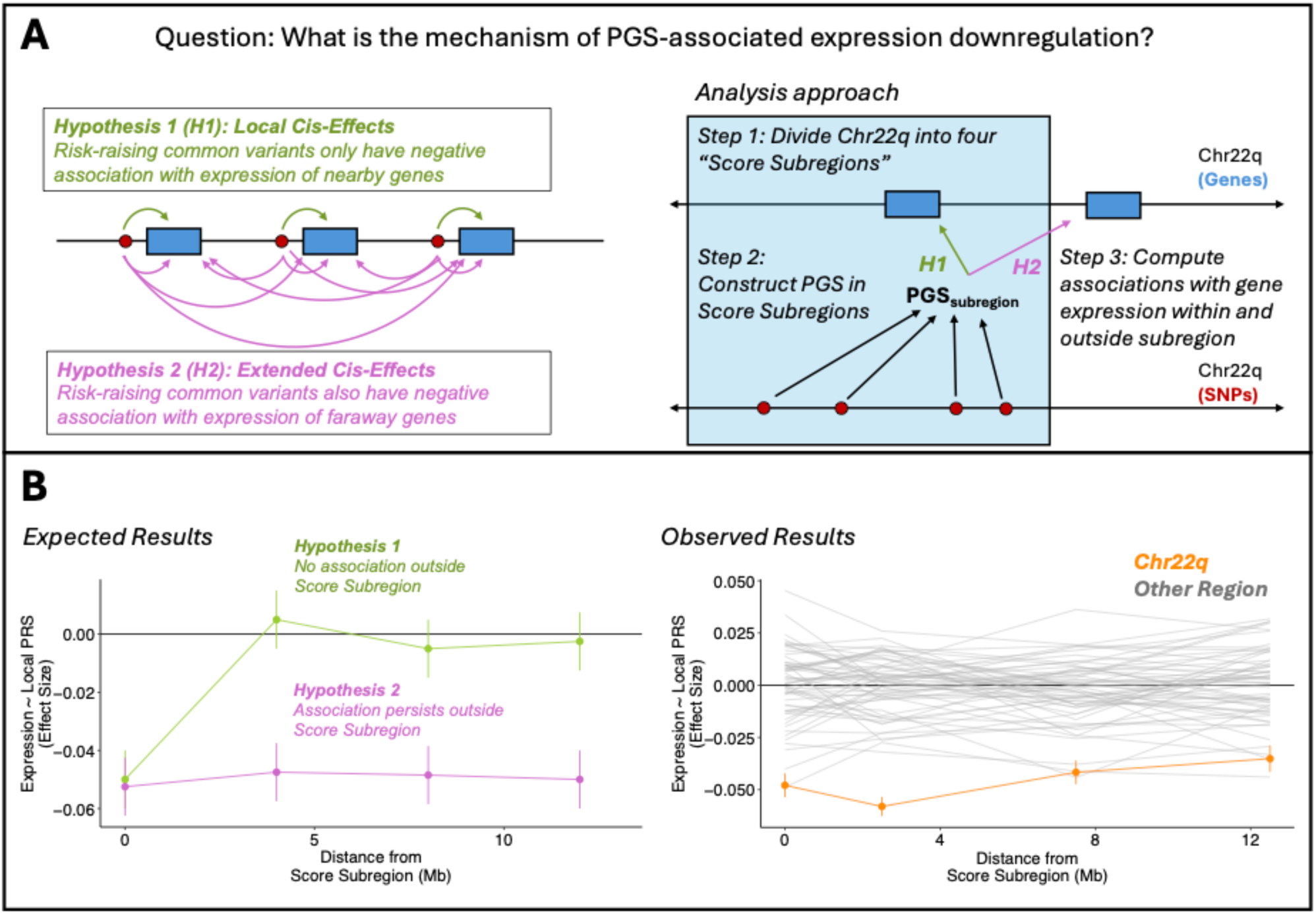
Distal expression associations of local polygenic scores. (A) Hypotheses to be examined. Hypothesis 1 entails several conventional regulatory interactions between SNPs and nearby genes, whereas Hypothesis 2 also includes distal SNP-gene interactions. Hypotheses are tested by evaluating association between sub-region polygenic score and genes inside and outside the sub-region. (B) Expected and observed results. Distal effects will only be observed if hypothesis 2 is true (pink points and line). For observed results, estimates are shown for chr22q (orange) and other similarly sized genomic regions (gray).

To assess these hypotheses, we divided chr22q, as well as other similarly sized genomic regions, into 8 Mb sub-regions, the smallest window size powered for statistical comparison (where power is driven by the number of variants in the polygenic score and the number of genes with expression measurement). For each subregion, we computed the local Aggregate-Psych polygenic score, and associated those scores and gene expression both *within* each sub-region and *across* adjacent sub-regions (**Figure 3A; Methods**). We found that polygenic risk for neuropsychiatric disorders was substantially associated with distal downregulation in gene expression at chr22q (**Figure 3B**). Specifically, expression downregulation was observed within the 8 Mb sub-regions (“proximal”; p < 0.001), but was also still detectable as far as 15 Mb away (“distal”; p < 0.001 for genes between 10 and 15 Mb away from the score sub-region). Common polygenic risk for neuropsychiatric disease has uncommonly long-range expression associations within chr22q, larger in magnitude than most areas of the genome (**Figure 3B**). We did, however, observe a distribution of long-range effects genome-wide, with 18 of 61 regions (of similar size to chr22q) showing evidence of at least one distal effect at a Bonferroni significant threshold. These 18 regions were nominally enriched for intrachromosomal 3D contact in fetal neocortex (p = 0.03, two sample t-test, **Supplementary Figure 5**; Won et al, 2016). This finding suggests that while we have focused here on chr22q, the types of transcriptomic effects conceptualized in Hypothesis 2, and observed in these analyses, may exist in many genomic neighborhoods, shaped by the genome’s 3D architecture or properties with which it is correlated.

### 22q11.2del downregulates gene expression across chr22q

After observing that local polygenic risk for neuropsychiatric diagnoses is associated with consistent downregulation of gene expression at chr22q, we asked whether the 22q11.2del, which is strongly associated with each of the aforementioned neuropsychiatric diagnoses, has similar functional consequences. We re-examined data from our previous functional genomic analysis of 22q11.2del in neuronal cultures (Nehme et al. 2022), collected two new datasets from these same cell lines, and additionally analyzed results from a published analysis of cerebral cortical organoids carrying 22q11.2del (Khan et al. 2020) (**Figure 4**). These studies span several experimental designs (i.e. case/control versus isogenic comparison), cellular states (i.e. earlier versus later neuronal cells), and model systems (i.e. 2D versus 3D culture). They are described in detail in **Methods**. In each dataset, we compared 22q11.2del and matched control samples in terms of chr22q gene expression, specifically chr22q genes *outside* the 22q11.2del region (ranging from 351 to 383 genes across datasets; **Figure 4A**).

**Figure 4:**
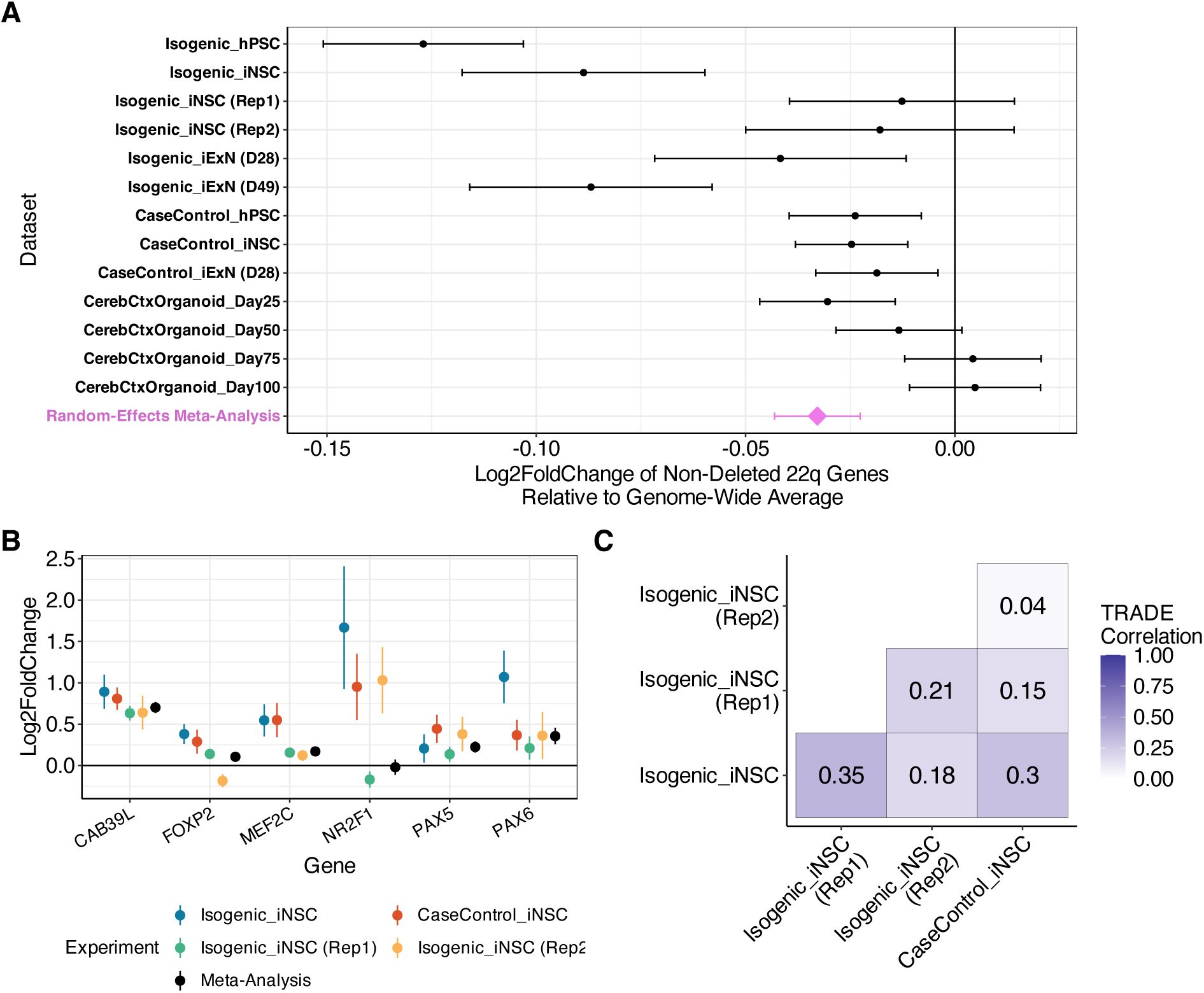
Convergent changes in chr22q11.2del. (A) Forest plot showing mean effects of 22q11.2del on non-deleted chr22q gene expression, including an estimate from a random-effects meta analysis. Error bars indicate standard errors. (B) Log2FoldChange estimates for previously identified key differentially expressed genes, across all induced neural stem cell (iNPC) datasets. Error bars indicate standard errors. (C) Estimated genome-wide log2FoldChange correlations for non-deleted genes between four iNPC 22q11.2del datasets.

Meta-analyzing across the collection of datasets, we found a significant downregulation of genes across chr22q due to the approximately 3 Mb 22q11.2del (p = 0.001; **Figure 4A**). This suggests that the common polygenic influences on neuropsychiatric diagnoses within chr22q operate in a functionally convergent manner with the 22q11.2del, the strongest genetic risk factor for schizophrenia at a population level. This is a second observance of chromosome-arm-wide convergence in the functional impact of common and rare variant risk factors for neuropsychiatric disorders. The first observation was made at chr16p, a region of the genome similarly harboring a classic neuropsychiatric disease CNV, and similarly characterized by exceptional intrachromosomal 3D contact (Weiner et al. 2022).

We also observed a significant heterogeneity estimate within the meta-analysis (p < 0.001). This analysis offered a rare opportunity to examine inter-experimental consistency of functional genomic experiments, an important question in V2F research. While some heterogeneity in broad expression downregulation by 22q11.2del is likely explained by experimental design, we also observed heterogeneity within similar experiments. Particularly surprising was the heterogeneity of effects between repeated examination of induced neuronal progenitor cells derived from the same cell line (i.e. between “Isogenic_iNPC”, “CaseControl_iNPC”, “Isogenic_iNPC (Rep1)”, and “Isogenic_iNPC (Rep2)” in **Figure 4A**), where inter-experimental variability is expected to be minimal. To better understand this variability, we further examined these four distinct datasets profiling gene expression in 22q11.2del-carrying induced neuronal progenitor cells from the same cell lines, compared with isogenic controls.

We found that the degree of inter-experiment consistency is dependent on gene expression effect size, with large expression changes being well-validated and highly replicable, while variability was more pronounced among smaller expression changes (**Figure 4B, C)**. In detail, we first assessed the replicability of key differentially expressed genes outside the 22q11.2del interval highlighted by Nehme et al (2022) **(Figure 4B**). For 4 of 6 genes, all experiments inferred effects in the same direction with a significant random-effects meta-analytic estimate (p < 0.001); for *FOXP2*, one estimate was in the opposite direction but with a significant meta-analytic estimate, and for *NR2F1*, the meta-analytic estimate was non-significant, likely owing to the low expression level (**Supplementary Figure 6**). We then estimated the correlation of effects across all genes (i.e. including many much smaller effects) using Transcriptome-wide Analysis of Differential Expression (TRADE; **Methods**) (Nadig et al. 2025). The correspondence of transcriptome-wide effects was more modest. For example, the inferred effect size correlations between the isogenic NPC datasets were 0.35, 0.18, and 0.21 (**Figure 5C**). These estimates suggest that, across the entire transcriptome, most differential expression effects were not shared between these experiments. Our findings suggest a complex landscape of robustness and heterogeneity in the transcriptional impact of 22q11.2del across experiments, where the largest effects are largely consistent, but the transcriptome-wide pattern of smaller effects is more variable.

## Discussion

We observed that polygenic risk for schizophrenia, autism, ADHD, and lower IQ behave unusually within chr22q, in that they result in surprisingly convergent expression downregulation across the vast genomic territory. We demonstrated these signals reflect associations between variants and expression of genes up to 15 Mb away, suggesting mechanisms other than classic *cis-*regulation. Lastly, we found that 22q11.2del exerts a similar downregulation on genes across chr22q. The common polygenic influences are also associated with increased co-occurrence of schizophrenia, autism, ADHD, and low IQ, which are strongly associated with 22q11.2del. These findings point towards phenotypic and functional convergence of rare and common genetic causes of neuropsychiatric disorders at chr22q.

These results expand several features of the observations first made in Weiner et al. 2022: that long range effects of common polygenic risk can be observed in multiple genomic regions; that those effects can occur over large genomic distances; that the effects can be shared across diagnoses and have unusually strong associations with lower cognitive ability. Future functional studies in larger samples, paired with expanding genetic association studies, will be able to identify the prevalence of similar patterns across the genome in an unbiased manner.

The largest remaining questions are those of mechanism, and potential special relevance to disorders of human behavior. How does common and rare genetic risk for neuropsychiatric disease exert these convergent functional effects across a chromosome arm? We hypothesize that these effects are mediated by low-dimensional patterns of 3D genome architecture, having demonstrated that both chr16p and chr22q are exceptional with respect to 3D intrachromosomal contact (as has been previously observed; Yaffe and Tanay 2011; Rao et al. 2014). However, regional 3D architecture is correlated with many genomic features, and to resolve this question, future work should attempt to disentangle possible explanations in larger datasets.

Why are these effects most obvious on chr16p and chr22q, the regions of the genome containing psychiatric genetics’ most studied CNVs? Is gross transcriptional dysregulation of brain expressed genes underlying the observed phenotypic convergence? These findings open new avenues for investigations of neuropsychiatric disorders, and greater synthesis of statistical and functional approaches to parse the genome. Central to these pursuits will be the collection of large scale resources with paired functional and genomic data in health and disease, as well as novel statistical methods to connect polygenic architecture to dispersed transcriptomic changes.

## Methods

### Region-level HiC Analysis

To compare the regulatory architecture of chr22q to other similarly sized regions, we constructed 61 33-Mb regions of the human genome, one of which represented chr22q (**Supplementary Table 1**). These regions were constructed to avoid centromeres and telomeres based on human genome build GRCh37.

To estimate the degree of intrachromosomal contact in each region in lymphoblastoid cell lines, we used previously described 1-Mb resolution HiC data from GM06990 (Lieberman-Aiden et al. 2009). For each region, we estimated the degree of intra-region contact as the mean of all elements in the 33 Mb x 33 Mb count matrix for that region. To estimate the degree of intrachromosomal contact in developing brain samples, we performed an identical analysis in a previously described 100kb-resolution HiC sequencing dataset in midgestational cortical plate (Won et al. 2016).

Degree of physical contact has been demonstrated to vary as a function of gene density and segmental duplication content (Weiner et al. 2022). In order to assess variation in physical content conditional on these genomic features, we regressed gene count and segmental duplication out of degree of physical contact. The scaled residuals from this model are used in the primary analysis in **Figure 1A**. To estimate segmental duplication content, we calculated the fraction of nucleotides for each region that overlapped at least one segmental duplication in the UCSC Genome Browser (Kent et al. 2002). We used BEDTools v2.30.0 to calculate segmental duplication coverage.

### Generation of local polygenic scores

We generated local polygenic risk scores for 8 complex traits and common diseases. For seven of these traits, we used publicly available summary statistics: schizophrenia (Trubetskoy et al. 2022) ADHD (Demontis et al. 2019), intelligence (Savage et al. 2018), height (Neale Lab), body mass index (Neale Lab), low-density lipoprotein (Neale Lab), and red blood cell count (Neale Lab). Neale lab summary statistics were downloaded from http://www.nealelab.is/uk-biobank/. For autism, we used an unpublished set of summary statistics from an in-progress update to the iPSYCH/Psychiatric Genomics Consortium autism GWAS (Grove et al. 2019). Briefly, this sample comprises 26,067 cases and 46,455 controls drawn from the Danish iPSYCH resource and the Psychiatric Genomics Consortium, which includes several cohorts from around the world.

We first used LD Score Regression (Bulik-Sullivan et al. 2015) to quality-control summary statistics, and estimate SNP-heritability. Then, to estimate polygenic score weights, we used the best linear unbiased predictor (BLUP) approach as implemented in LDPred2-Inf (Privé, Arbel, and Vilhjálmsson 2021). To calculate the local polygenic score, we filtered these weights to SNPs residing within each of the 33-Mb genomic regions described above. We used PLINK2 (Chang et al. 2015) to compute polygenic scores from genotypes and LDPred2-inf derived weights.

### Single-nucleus RNA sequencing of postmortem human brain

We analyzed a resource of paired genotypic and single-nucleus RNA-seq data from the dorsolateral prefrontal cortex (DLPFC) of post-mortem human brain samples from the HBTRC/NIH NeuroBioBank; this dataset is described in detail in the primary manuscript describing it (Ling et al. 2024). Briefly, we developed a workflow for generating and analyzing pools of nuclei from DLPC from 20 donors per pool. We started by dissecting tissue from each donor, ensuring that we obtained a similar mass of tissue from each specimen while sampling all cortical layers. The frozen tissue samples were then pooled for simultaneous isolation of nuclei, and subsequent steps (nuclear isolation, encapsulation in droplets and preparation of snRNA-seq libraries) used all donors pooled together. This “dropulation” workflow minimizes experimental variability, including effects of messenger RNA ascertainment and cell-free ambient RNA. We reassigned nuclei to donor-of-origin using combinations of hundreds of transcribed SNPs that disambiguated individual genotypes. We performed global clustering and identification of marker genes to assign nuclei to seven major cell classes (glutamatergic neurons, GABA-ergic neurons, astrocytes, oligodendrocytes, polydendrocytes, microglia, and endothelia). Median cell-type proportions were: 48% glutamatergic neurons, 19% GABA-ergic neurons, 14% astrocytes, 8% oligodendrocytes, 5% polydendrocytes, 2% microglia, and 1% endothelia. The cell type-specific gene-by-donor expression matrices were normalized and variance-stabilized using the *scTransform* utility (Hafemeister and Satija 2019). As previously described (Weiner et al. 2022), we used PCA to identify European ancestry samples and exclude samples with mean expression >3 s.d. from the cohort mean, yielding a final sample of 122 individuals. Notably, this cohort includes individuals with schizophrenia (n = 58); diagnosis was included as a covariate in all models presented.

### Correcting for gene density

We confirmed that unusual PGS/expression relationships at chr22q are not driven by unusual gene density in this region. In particular, we computed the number of genes in each region by assessing whether the midpoint of each gene lies within the region boundaries, as previously described (Weiner et al. 2022). We additionally computed the number of brain-expressed genes by identifying the 10% of genes with cortex-specific expression in bulk RNA-seq data from the GTEX project (GTEx Consortium 2013), by Z-scoring expression medians across tissues (excluding brain tissues other than “Brain (Cortex)”, and retaining the top 10% of genes by “Brain (Cortex)” Z-score. For each neuropsychiatric trait, we observed that while chr22q is exceptional with respect to gene and brain gene density across regions, that this does not explain the average negative effect of local PGS on expression in this region. (**Supplementary Figure 7**)

### Bulk-RNA sequencing of postmortem human brain

We analyzed a resource of paired genotypic and bulk RNA-seq from dorsolateral prefrontal cortex (DLPFC) samples from the CommonMind consortium (Hoffman et al. 2019). Generation of the expression count matrices is described extensively in the primary publication for this resource (Hoffman et al. 2019). Within this dataset, we restricted analysis to donors from the NIMH Human Brain Collection Core (HBCC) and the University of Pittsburgh (PITT) biobanks due to previous analysis suggesting high concordance with the snRNA-seq resource described above. We processed these data with *scTransform* with similar parameters as our snRNA-seq analysis.

As previously described (Weiner et al. 2022), we used PCA to identify European-(N = 302) and African-ancestry (N = 206) samples, and analyzed each population separately. Notably, this cohort includes individuals with diagnoses of schizophrenia (n = 69) and bipolar disorder (n = 89)

### Local genetic correlation analysis with rho-HESS

We estimated local genetic correlations with rho-HESS (Shi et al. 2017). For this analysis, we used an LD reference panel from the 1000 Genomes Project and a genome partition composed of 1453 approximately LD-independent blocks (Berisa and Pickrell 2016). To estimate local genetic correlations for large genomic regions, we performed precision weighted meta analysis for heritabilities and genetic covariance across all LD-blocks in the large genomic region. To estimate the standard error of the local genetic correlation for large genomic regions accounting for uncertainty in local heritability and genetic covariance, we used the delta method.

### Phenotypic associations in the UK Biobank

The UK Biobank is a longitudinal study of roughly 500,000 volunteers who were recruited between 2006 and 2010 when they were 40–69 years-old at enrollment. The UK Biobank version 3 imputed genetics data was used in our analyses. We included unrelated individuals with low autosomal missingness rates used for PCA who were predominantly estimated European genetic ancestry based on analysis of the top six principal components (PCs) (Bycroft et al. 2018). After excluding individuals who withdrew from UK Biobank participation and sample quality control, there were 361,086 individuals included in our analyses. We restricted variants with minor allele frequency > 0.05, missing rate < 0.05, and Hardy-Weinberg Equilibrium (HWE) p-value > 1 x 10^-10^ to be included in the analyses. We generated local polygenic scores for 5 traits, including autism, schizophrenia, ADHD, intelligence and height with the same approach as previously described and standardized the polygenic scores. For the phenotypes, we used height (field ID: 50, N = 360,263), fluid intelligence score (field ID: 20016, N = 117,093) and education attainment (college or university education or above, field ID 6138, N = 360,558). The analytical sample sizes were different because of the missingness of the phenotypes. Using standardized polygenic scores and phenotypes, we ran linear regression models for all the combinations between local polygenic scores and phenotypes, adjusting for age, sex, interaction term of age and sex, squared age and top 20 PCs.

### Distal prediction with local polygenic scores

To estimate associations between local polygenic scores and distal genes, we first subdivided the 61 33-Mb region (“parent” region) from our main analyses into four ∼8Mb sub-region (“child” region) each. For each child subregion, we computed local polygenic scores as described above.

Then, in glutamatergic neurons from the same single-cell post-mortem human brain dataset above, we used linear models to estimate associations between local polygenic scores and normalized/variance-stabilized expression of genes across the entire parent region. We examined average polygenic score/expression associations within each child region, including the child subregion used to calculate the polygenic score (i.e. “proximal” effects), and the adjacent child subregions within the same parent region (i.e. “distal” effects).

### Differential expression analyses in isogenic cell lines

We reanalyzed previously described bulk-RNA sequencing data in two 22q11.2del-carrying cell lines and isogenic controls, in four different *in vitro* cell types: human pluripotent stem cell (hPSC), induced neural progenitor cell (iNSC), and two induced excitatory neuron stages (iExN-Day28 and iExN-Day49). Briefly, these cell lines were generated from H1 hESCs with CRISPR-Cas9 sgRNAs targeted towards LCR-A and LCR-D, the most common breakpoints for recurrent 22q11.2del. The protocol for generation and differentiation of these cell lines was previously described in detail (Nehme et al. 2022).

For each cell-type, we analyzed deletion and isogenic control data from two cell lines, with 3 replicates each (e.g. four cell lines * 3 replicates each = 12 total samples). We performed differential expression analysis with a regularized negative binomial generalized linear model, implemented in DESeq2 (Love, Huber, and Anders 2014). Briefly, DESeq2 models the mean-variance relationship across genes to regularize overdispersion estimates, and uses these regularized estimates to estimate log2FCs. For normalization, we used the default DESeq2 median-of-ratios approach, which estimates a common size factor across all genes. We used surrogate variable analysis (Leek and Storey 2007) to control for unobserved confounding variables.

For each cell type, we used the following regression equation:

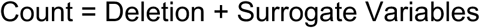

### Differential expression in case/control cohorts

We used a similar analytic strategy to assess differential expression in a case-control sample collection from the same previous study (Nehme et al. 2022). For this analysis, we had access to data from three cell types: hPSC, iNPC, and iExN (e.g. without a second iExN stage, as in the isogenic analysis). For each cell type, we had data from 28 control individuals and 20 individuals carrying 22q11.2del, for a total of 144 samples. For this analysis, we also used DESeq2 and SVA to perform differential expression analysis, with identical regression equations to those for the isogenic analysis.

### Differential expression in organoids

We analyzed gene-wise differential expression summary statistics from patient– and control-derived cerebral cortical organoids (Khan et al. 2020), that were shared by the authors of the primary manuscript, where experimental and analytic details can be found.

### Generation of new expression data from isogenic cell lines

Human ESCs and iPSCs were maintained on plates coated with geltrex (life technologies, A1413301) in StemFlex media (Gibco, A3349401) and passaged with accutase (Gibco, A11105). All cell cultures were maintained at 37 °C, 5% CO2.

hPSCs were differentiated into cortical glutamatergic neurons as previously described (Nehme et al. 2018). Our protocol differs from previous Ngn2-driven protocols (Theka et al. 2013; Busskamp et al. 2014) through inclusion of developmental patterning alongside Ngn2 programming. This paradigm generates post-mitotic excitatory cortical neurons that are highly homogeneous in terms of cell type compared to most differentiation paradigms which yield heterogeneous cell types (Chambers et al. 2009). At 4 days post induction, cells are co-cultured with mouse glia to promote neuronal maturation and synaptic connectivity (Eroglu and Barres 2010; Pfrieger 2009; Pietiläinen et al. 2023).

For the first dataset collected (“Isogenic_iNSC_Rep1”), the neuronal mono-cultures and co-cultures underwent bulk RNA sequencing at the Broad Genomics Platform according to the Smartseq2 workflow. For the second dataset collected (“Isogenic_iNSC_Rep2”), samples were sequenced on an Illumina NextSeq500 instrument. For both of these datasets, read alignment to human genome build hg19 was performed with STAR (Dobin et al. 2013), and transcript abundances were estimated with RSEM (Li and Dewey 2011).

### Estimating effect size correlations with TRADE

We estimated the correlation between differential expression effect sizes across experiments with TRADE (Nadig et al. 2024). Briefly, TRADE fits a flexible mixture model to the joint distribution of observed log2FoldChanges and standard errors, to provide unbiased estimates of the effect size correlation. For these analyses, which focused on datasets with 22q11.2del samples, we excluded genes within the deletion interval.

## Data availability

Raw sequencing data for novel datasets will be uploaded to GEO prior to publication; data can be shared with reviewers upon reviewer request.

