## Supplementary Figures for "Functional consequences of genetic risk for neuropsychiatric conditions at chr22q"

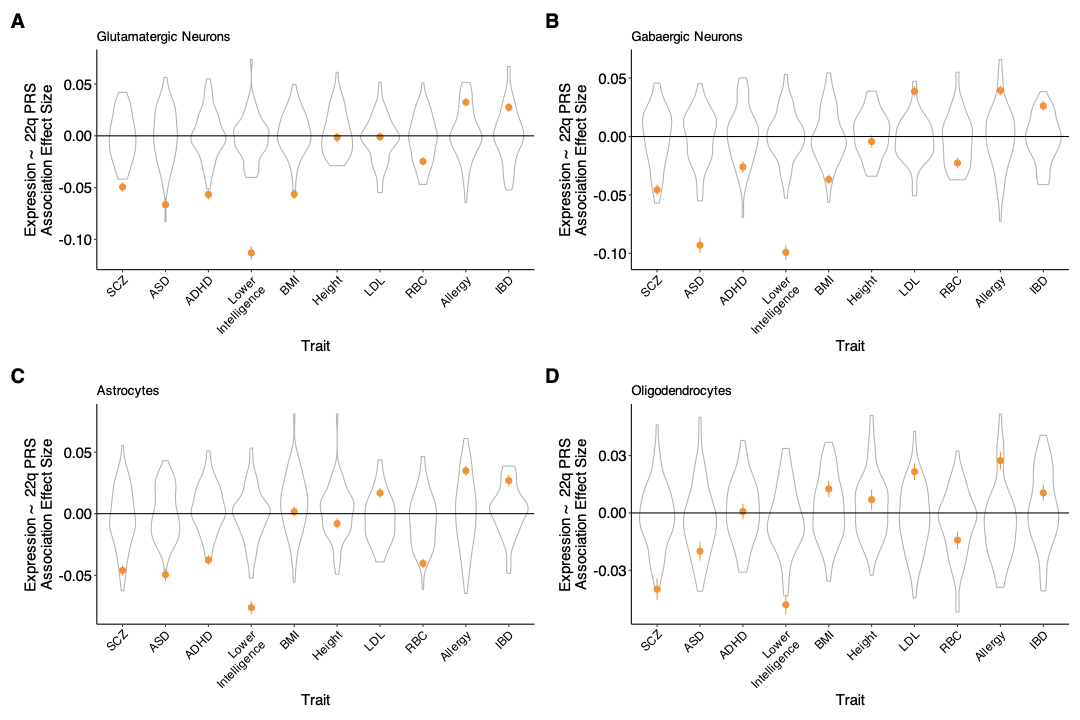

**Supplementary Figure 1. Associations between local PGS and gene expression across four brain cell types**. Orange points indicate association between chr22q local PGS and average expression of chr22q genes. Grey indicates distribution of local PGS/expression associations across 60 other similarly sized genomic regions. Results are shown for (A) glutamatergic neurons, (B) gabaergic neurons, (C) astrocytes, and (D) oligodendrocytes.

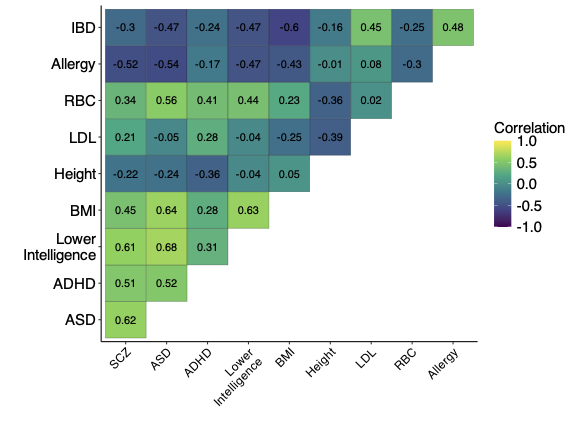

**Supplementary Figure 2. Correspondence of local PGS/expression associations at chr22q across traits.** Each correlation value represents the correlation of gene-wise local PGS/expression regression effect sizes at chr22q.

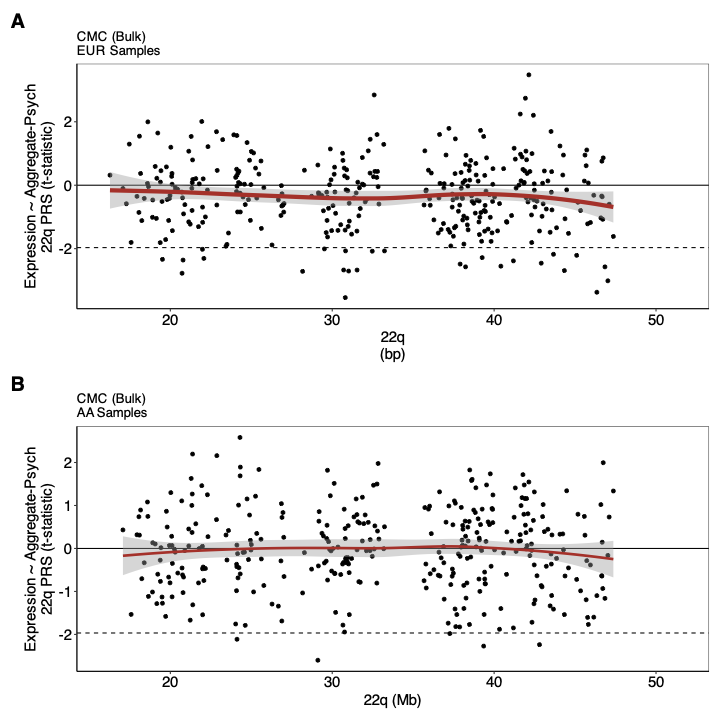

**Supplementary Figure 3. Local PGS/expression relationship in bulk post-mortem RNA-seq data from the CommonMind consortium**. Association t-statistics between local chr22q Aggregate-Psych PGS and expression of genes on chr22q in (A) European and (B) African ancestry individuals from the CommonMind consortium.

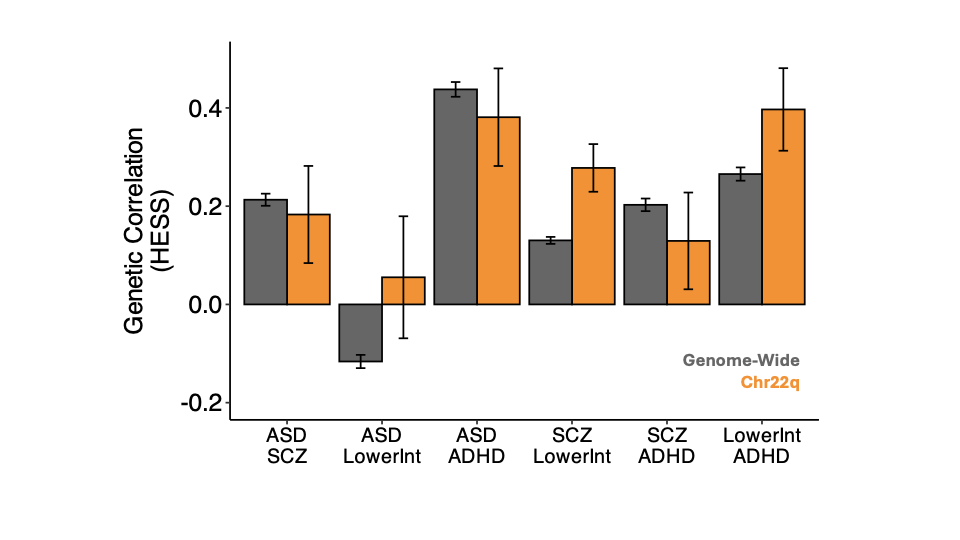

**Supplementary Figure 4.** rho-HESS estimates of genome-wide and chr22q local genetic correlations between neuropsychiatric traits and diagnoses. Error bars indicate standard errors.

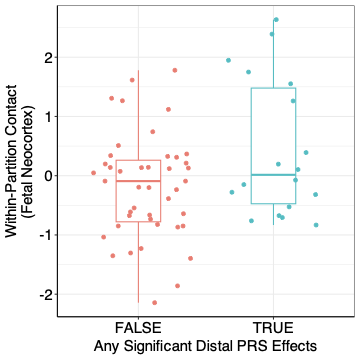

**Supplementary Figure 5. Intrachromosomal physical contact estimates for large genomic regions, stratified by presence of distal expression associations**. Each point represents a 33 Mb genomic region, x axis values indicate the presence or absence of distal PGS/expression relationships, and y axis values represent z-scored degree of intrachromosomal contact estimated from a developing cortical plate HiC dataset (Won et al, 2016)

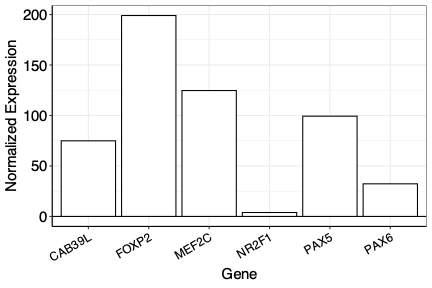

**Supplementary Figure 6**. Normalized expression levels of key differentially expressed genes, averaged across iNSC datasets, including both 22q11.2del and isogenic control cells.

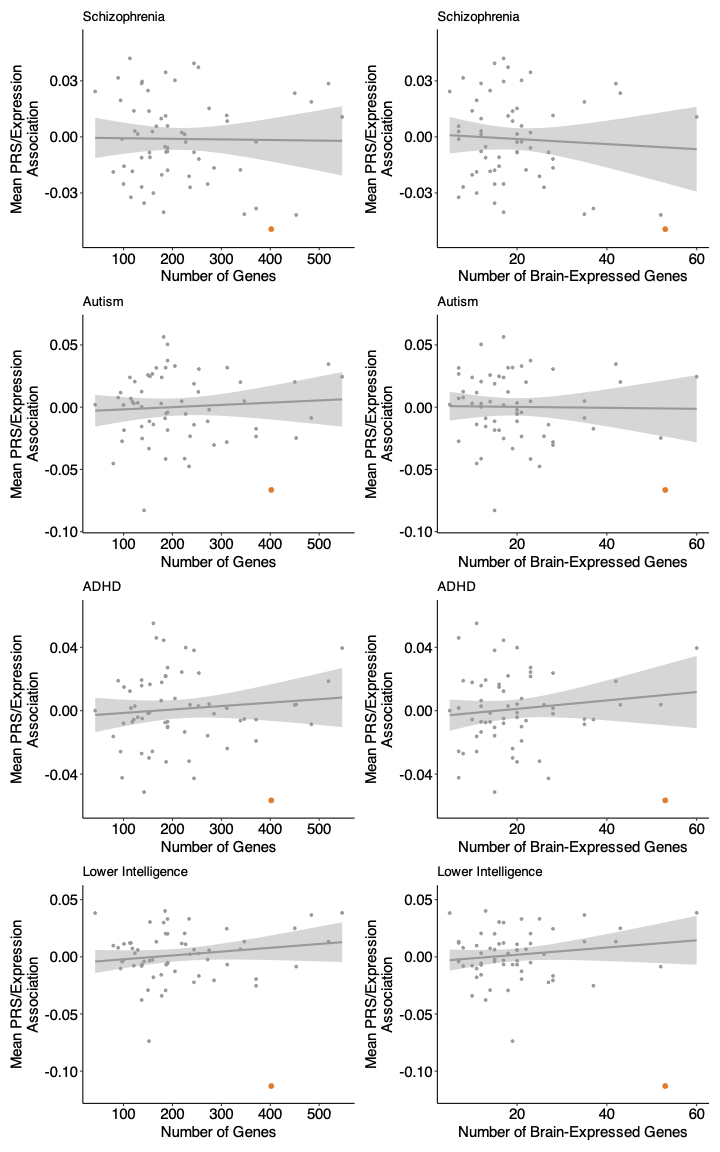

**Supplementary Figure 7**: **Average local PGS/expression relationship across 33-Mb regions, compared with gene density estimates**. Plots in the left column demonstrate no relationship between gene density and average PGS/expression relationship. Plots in the right column demonstrate no relationship between brain gene density (**Methods**) and average PGS/expression relationship. Chr22q is shown in orange.
